# Adverse events reported during weekly isoniazid-rifapentine (3HP) tuberculosis preventive treatment among people living with HIV in Uganda

**DOI:** 10.1101/2024.08.08.24311632

**Authors:** Jillian L. Kadota, Allan Musinguzi, Hélène E. Aschmann, Lydia Akello, Fred Welishe, Jane Nakimuli, Christopher A. Berger, Noah Kiwanuka, Patrick PJ Phillips, Achilles Katamba, David W. Dowdy, Adithya Cattamanchi, Fred C. Semitala

**Affiliations:** Division of Pulmonary and Critical Care Medicine, University of California San Francisco, San Francisco CA USA; Center for Tuberculosis, University of California San Francisco, San Francisco CA USA; Infectious Diseases Research Collaboration, Kampala, Uganda; Department of Epidemiology and Biostatistics, School of Public Health, Makerere University College of Health Sciences, Kampala, Uganda; Clinical Epidemiology & Biostatistics Unit, Department of Medicine, Makerere University College of Health Sciences, Kampala, Uganda; Uganda Tuberculosis Implementation Research Consortium, Kampala, Uganda; Department of Epidemiology, Johns Hopkins Bloomberg School of Public Health, Baltimore, MD, USA; Department of Medicine, Makerere University College of Health Sciences, Kampala, Uganda; Division of Pulmonary Diseases and Critical Care Medicine, University of California Irvine, Orange CA USA; Makerere University Joint AIDS Program, Kampala, Uganda

## Abstract

**Background:** Short-course tuberculosis (TB) prevention regimens, including 12 weeks of isoniazid and rifapentine (3HP), are increasingly used in high TB-burden countries. Despite established safety and tolerability in efficacy trials, 3HP-related adverse events (AE) could differ in routine settings. Real-world data on AE type, frequency, and timing is crucial for health systems considering 3HP programmatic scale-up.

**Methods:** We reviewed AEs among people living with HIV (PLHIV) participating in a pragmatic implementation trial of facilitated 3HP taken by directly observed therapy (DOT) or self-administered therapy (SAT) in Kampala, Uganda, and classified them using the Common Terminology Criteria for Adverse Events. We assessed AE timing and summarized related clinical actions including lab tests, diagnoses made, medications prescribed, and treatment interruptions.

**Results:** Among 1655 PLHIV treated between July 2020-September 2022, 270 (16.3%) reported 451 events; main issues included general (7%), nervous system (6%), musculoskeletal (5%), gastrointestinal (5%), and dermatologic (3%) disorders. Most (61%) occurred within 6 weeks of initiating 3HP. Among those with events, 211 (78%) required further clinician evaluation, 202 (75%) required laboratory testing, 102 (38%) had medications prescribed, 40 (15%) had treatment paused, and 14 (5%) discontinued 3HP. Women, those multidimensionally impoverished, and DOT recipients were more likely to report an AE. SAT users and later enrollees were more likely to have 3HP interrupted or stopped due to an AE.

**Conclusions:** In a routine setting, 3HP was safe with 16% of PLHIV reporting AEs and only 3% requiring temporary or permanent treatment interruption. These findings support 3HP expansion in routine HIV/AIDS care settings for TB prevention.

**Summary:** 3HP is being rolled out for TB prevention in high burden countries. We describe the incidence and clinical management of adverse events in a real-world clinical setting among people living with HIV participating in a pragmatic implementation study in Uganda.

## BACKGROUND

Prevention of tuberculosis (TB) is an urgent priority for global TB programs (1,2) and scale-up of TB preventive treatment (TPT) in high TB burden countries is critical for achieving ambitious TB elimination goals by 2030. Short-course regimens, such as 12 weekly doses of isoniazid and rifapentine (3HP), are now recommended as an option for TB prevention based on several trials demonstrating efficacy, safety, and improved completion relative to the traditional standard of six to nine months of daily isoniazid (3–6). However, data on the safety of 3HP in routine care settings remain sparse and are needed to help policymakers better anticipate the resources required for adverse event management as 3HP is rolled out for TB prevention.

Here, we describe adverse events reported in the *3HP Options Trial*, a pragmatic implementation trial that delivered 3HP to adult PLHIV accessing routine clinical HIV/AIDS care in Kampala, Uganda via facilitated directly observed therapy (DOT), facilitated self-administered therapy (SAT) or participant choice between facilitated DOT and SAT. In this pragmatic trial, routine health workers were responsible for monitoring, evaluating, and providing care to participants receiving 3HP treatment. We previously reported that treatment completion was high overall (94%), and similar across trial arms (95%, 92% and 94% for facilitated DOT, facilitated SAT and Choice, respectively) (7). Our objective here was to characterize the frequency, type and severity of adverse events, and secondarily to identify demographic and clinical characteristics associated with reported adverse events and with 3HP treatment interruption or discontinuation.

## METHODS

### Study design and population

The study design and population, including eligibility criteria, are described in detail elsewhere (7,8). Briefly, the *3HP Options Trial* was a pragmatic trial of 3HP delivery strategies among adult (≥18 years) PLHIV accessing care at the Mulago Immune Suppression Syndrome (IS;) clinic, the largest HIV/AIDS clinic in Uganda. Eligible PLHIV were randomized to one of three delivery strategies: facilitated directly observed therapy (DOT), facilitated self-administered therapy (SAT), or participant choice between facilitated DOT and SAT. After taking the first dose in person, participants taking 3HP via facilitated DOT returned to the clinic weekly for the remaining 11 doses, whereas those taking 3HP via facilitated SAT had scheduled clinic visits for in-person dosing at weeks 6 and 12 only.

### Adverse events reporting and assessment

Prior to 3HP dosing at scheduled clinic visits, a Mulago ISS clinic pharmacy technician used a standardized form to screen participants for the presence/absence of any side effect and if present to screen for each of the following commonly reported side effects of 3HP treatment: 1) loss of appetite, 2) nausea or vomiting, 3) yellow eyes or skin, 4) abdominal pain, 5) diarrhea, 6) rash/hives, 7) fever or chills, 8) dizziness/fainting, 9) numbness or tingling, 10) headache, 11) joint pain, 12) itching, 13) weakness, or 14) other (with the option to specify). Following self-administered doses, participants taking 3HP by SAT could report that they were feeling unwell via weekly two-way toll-free interactive voice response (IVR) phone calls sent by a digital adherence monitoring platform used for the study (99DOTS, Everwell Health Solutions, Bengaluru, India). Mulago ISS clinic pharmacy technicians made phone calls to participants who reported feeling unwell and screened for side effects using the same standardized form used during in-person visits. In addition, all participants were instructed that they could visit or call the clinic at any time during open hours if they felt unwell, without the need to wait for their next scheduled appointment. Thus, participants could report adverse events at scheduled clinic visits for in-person dosing, IVR phone calls (SAT only), or through unscheduled phone calls and clinic visits.

Mulago ISS clinic pharmacists recorded the presence or absence of each common side effect using the standardized form. The form also included fields to specify if the patient was referred for further evaluation by a clinician, if or which laboratory tests were ordered, results of any laboratory testing, medications prescribed because of adverse event(s) reported, and details about treatment interruption or permanent discontinuation. Of note, Mulago ISS clinic pharmacists made all decisions to refer participants with suspected adverse events to a routine (*i*.*e*., non-study) clinician who then managed all activities related to adverse events assessment, including additional clinical or laboratory evaluation and decisions to hold or discontinue 3HP treatment.

### Statistical analysis

At the conclusion of participant follow-up, two analysts (JLK and HEA) independently re-coded all adverse events initially recorded as “other” (*e*.*g*., those not in pre-defined categories and/or those with different spellings). The same analysts then independently re-categorized all adverse events using the Common Terminology Criteria for Adverse Events (CTCAE), version 5.0 (9). Two research staff with medical training (AM and LA) adjudicated disagreements in categorizations.

Time to adverse event onset was calculated by taking the difference between the date of first report of the adverse event and the date of 3HP initiation. We quantified adverse events that resulted in referrals, additional lab tests ordered, diagnoses made, and medications prescribed using counts and percentages, and compared across study arms using Fisher’s exact tests. Finally, we used multivariable logistic regression to explore the clinical and sociodemographic variables associated with reporting any adverse event and discontinuing treatment because of an adverse event. Variables included in final multivariate models were those determined *a priori* based on relationships identified in the literature (10,11).

The trial is registered at ClinicalTrials.gov (NCT03934931) and was approved by the ethical committees at the University of California San Francisco, Makerere University College of Health Sciences School of Public Health, and the Uganda National Council for Science and Technology. All participants gave written informed consent for study participation.

## RESULTS

Between July 13, 2020, and July 8, 2022, we enrolled 1,655 eligible PLHIV. Follow-up continued until September 29, 2022. In the Choice arm, 370 participants (67.0%) initially preferred facilitated DOT; thus, 921 participants received 3HP via facilitated DOT, and 734 participants via facilitated SAT. Median participant age was 42 years (interquartile range [IQR]: 36-48); 1,122 (67.8%) were female; and median time on ART was 9.0 years (IQR: 5.6-12.5), with no differences by treatment strategy (**Table 1**).

**Table 1.** Participant characteristics comparing those taking 3HP via directly observed therapy versus self-administered therapy (n=1,655).^1^.

|  | **DOT**<br>**(n=921)** | **SAT**<br>**(n=734)** |
| --- | --- | --- |
| **Age** | 41.9 (9.4) | 42.6 (9.2) |
| **Female sex** | 614 (66.7) | 508 (69.2) |
| **Education** |  |  |
| None | 71 (7.7) | 68 (9.3) |
| Primary | 452 (49.1) | 330 (45.0) |
| Secondary | 335 (36.4) | 265 (36.1) |
| Tertiary/Vocational | 37 (4.0) | 48 (6.5) |
| University/Graduate School | 26 (2.8) | 23 (3.1) |
| **Employment status<sup>2</sup>** |  |  |
| Unemployed | 129 (14.0) | 96 (13.1) |
| Hired worker | 105 (11.4) | 99 (13.5) |
| Self-employed worker | 445 (48.3) | 372 (50.7) |
| Temporary/informal worker | 233 (25.3) | 163 (22.2) |
| Other | 3 (0.3) | 2 (0.3) |
| **Multidimensional Poverty Index<sup>3</sup>** |  |  |
| Not vulnerable to multidimensional poverty | 378 (41.0) | 341 (46.5) |
| Vulnerable to multidimensional poverty | 332 (36.1) | 251 (34.2) |
| Multidimensionally poor | 167 (18.1) | 118 (16.1) |
| Severely multidimensionally poor | 44 (4.8) | 24 (3.3) |
| **Reported an adverse event while taking 3HP** | 175 (19.0) | 95 (12.9) |
| **Prior TB infection** | 165 (17.9) | 136 (18.5) |
| **Time (years) on ART** | 8.8 (4.5) | 8.9 (4.4) |
| **ART Regimen** |  |  |
| Dolutegravir + Lamivudine + Tenofovir | 804 (87.3) | 647 (88.2) |
| Tenofovir + Lamivudine + Efavirenz | 65 (7.1) | 46 (6.3) |
| Other<sup>4</sup> | 52 (5.6) | 41 (5.6) |
| **Viral load <1000 copies/mL<sup>5</sup>** | 906 (98.4) | 727 (99.1) |
| **Body Mass Index (kg/m<sup>2</sup>)** | 25.9 (5.8) | 26.3 (5.4) |

ART=antiretroviral therapy; DOT=directly observed therapy; SAT=self-administered therapy; TB=tuberculosis
1. Data are n (%) or mean and standard deviation (SD).
2. n=8 missing.
3. The global multidimensional poverty index (MPI) examines deprivations across 10 indicators in dimensions of health, education, and standards of living, with those deprived in one-third or more of the 10 indicators counted as being multidimensionally poor. MPI scores can range from 0 to 1 and are classified as not vulnerable to multidimensional poverty (MPI score: 0-0.19), vulnerable to multidimensional poverty (MPI score: 0.20-0.32), multidimensionally poor (MPI score: 0.33-0.49) and severely multidimensionally poor (MPI score: ≥0.50).
4. Other regimens included Abacavir + Lamivudine + Dolutegravir (n=82), Lamivudine + Zidovudine + Dolutegravir (n=8), Lamivudine and Zidovudine 150mg/300mg tablet (n=1), Abacavir + Lamivudine + Efavirenz (n=1).
5. Suppressed defined as <1000 copies/mL according to Ugandan guidelines; n=2 missing.

### Adverse event frequency, typology and timing

Overall, 270 (16.3%) participants reported at least one adverse event over the course of 3HP treatment (**Figure 1**), including 175 (19.0%) people receiving 3HP via DOT and 95 (12.9%) receiving 3HP via SAT (**Table 1**). Participants in the DOT arm, those who were female, or multidimensionally poor people were more likely to report any adverse event, associations confirmed in multivariable logistic regression models (adjusted odds ratios [aORs]=1.59, 95% confidence interval (CI): 1.21, 2.09; p=0.001 for DOT; 1.61, 95% CI: 1.05-2.01; p=0.02 for female sex; and 1.40, 95% CI: 1.00, 1.96; p=0.05; for poverty or severe multidimensional poverty) (**Supplement Table 1**).

**Figure 1.**
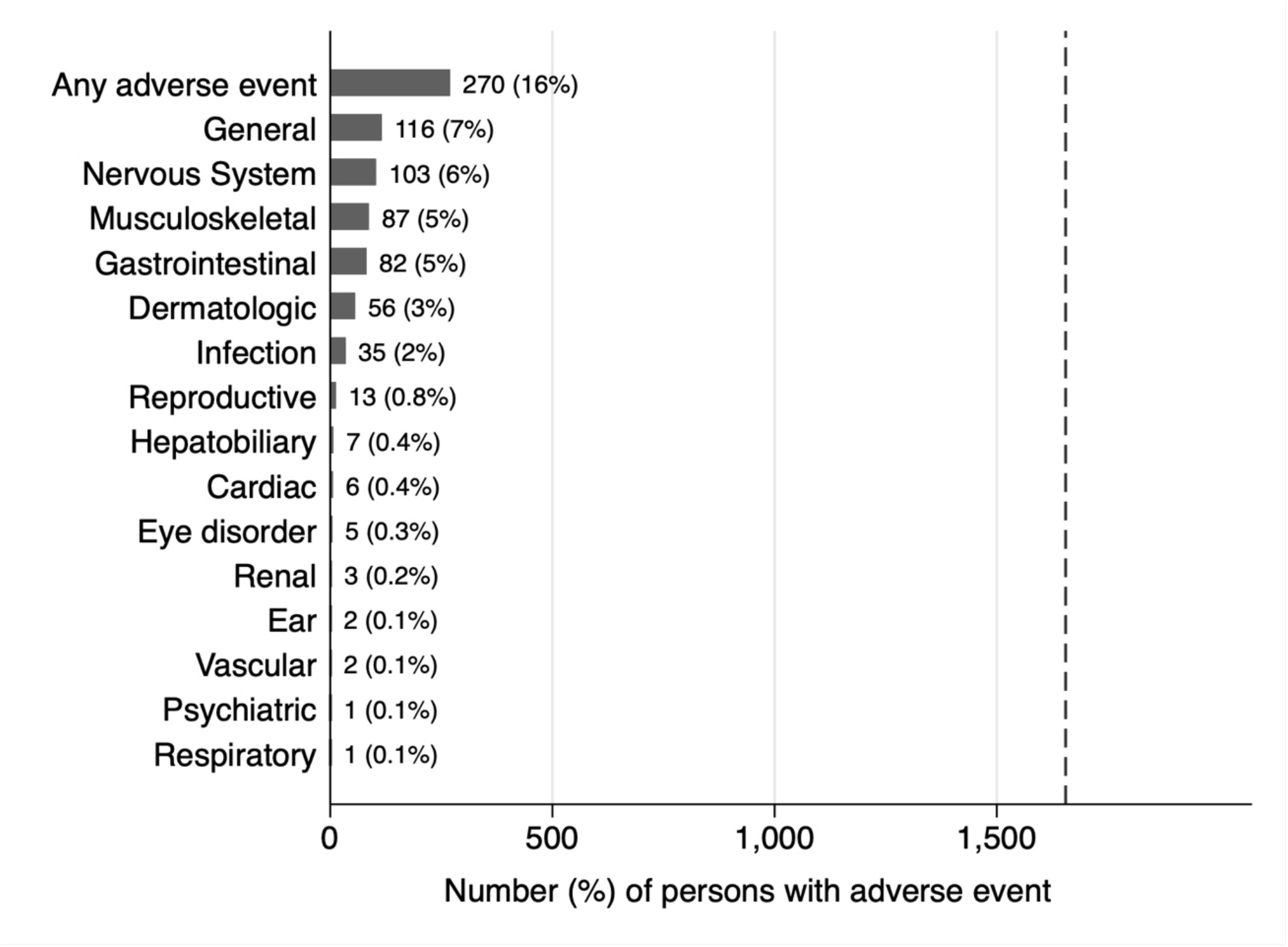
Frequency of reported adverse events during 3HP treatment according to Common Terminology Criteria for Adverse Events (version 5.0) categories. Shown are the number and percentage of participating PLHIV taking 3HP for prevention of tuberculosis who reported adverse events in each category. The total number of participants assessed for adverse events in the 3HP Options Trial was 1,655, as indicated by the dashed vertical line.

The most frequently reported adverse events (reclassified according to CTCAE, **Supplement Table 2**) were general disorders such as fever or flulike illness (n=116 PLHIV, 7% of all study participants), nervous system disorders (n=103, 6%), musculoskeletal disorders (n=87, 5%), gastrointestinal disorders (n=82, 5%), or dermatologic disorders (n=56, 3%; **Figure 1**). Hepatobiliary disorders were infrequent, with only seven participants (0.4%) reporting related adverse events.

Among participants who had an adverse event, the median time to an adverse event was 35 days (IQR: 14-49). There was a significant difference in the survival curves comparing time to an adverse event between participants receiving 3HP via DOT vs. SAT, with people receiving 3HP by SAT less likely to report an adverse event (log-rank p=0.001; **Figure 2**).

**Figure 2.**
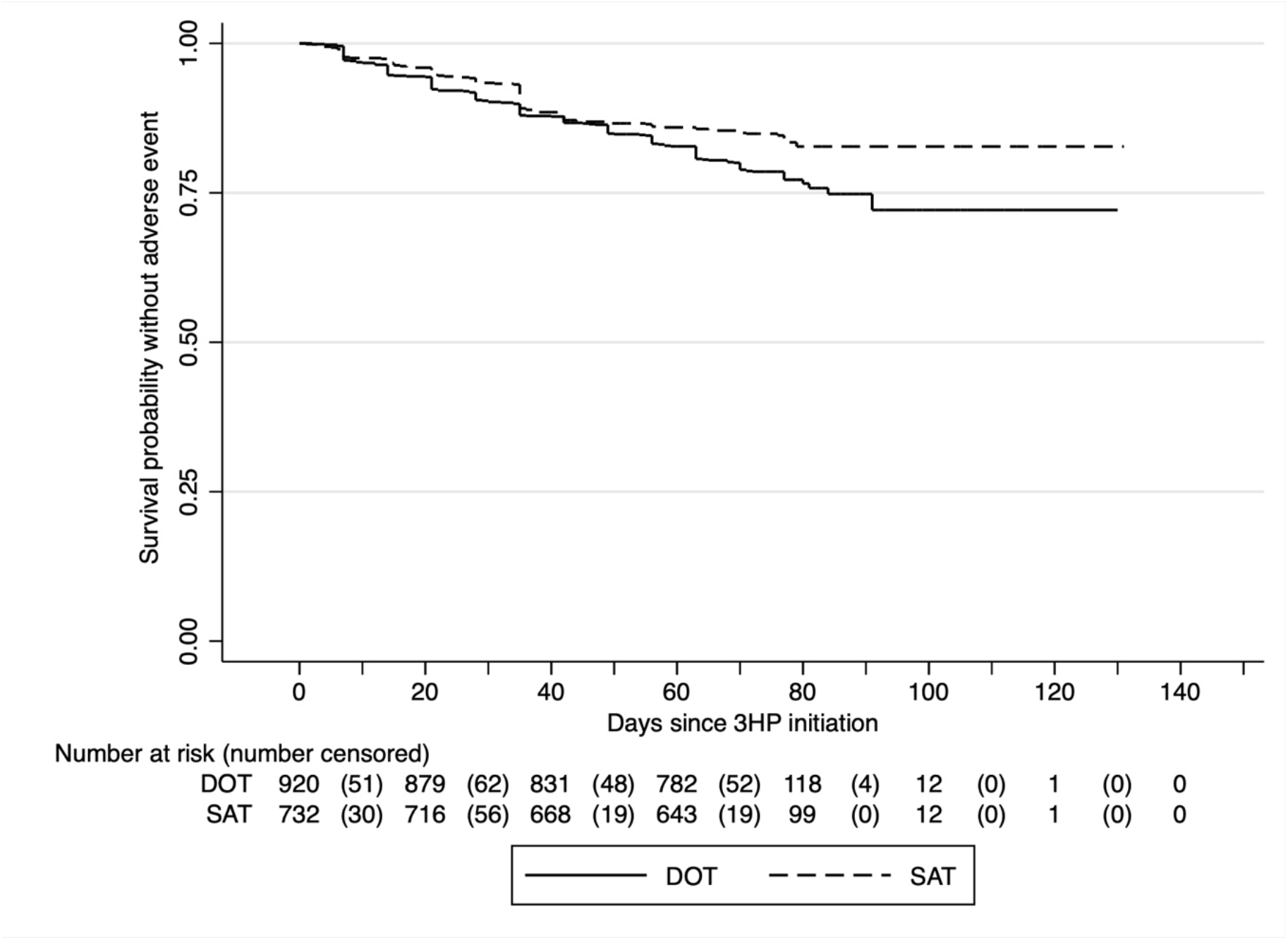
Time to first adverse event experience stratified by 3HP delivery strategy (facilitated DOT vs. facilitated SAT). DOT=directly observed therapy; SAT=self-administered therapy The solid horizontal stepped line represents the cumulative probability of not have an adverse event among participants receiving 3HP via facilitated directly observed therapy (DOT), and the dashed horizontal stepped line represents those receiving 3HP via facilitated self-administered therapy (SAT). The numbers outside of parentheses at the bottom of the figure represent the total number of participants at risk, while the numbers in parentheses represent the number censored at the time interval indicated.

### Additional health care utilization for adverse events

Among those reporting adverse events, 211 (78.1%) were referred to a clinician for further evaluation, 202 (74.8%) required laboratory testing, and 102 (37.8%) had medication(s) prescribed. The most common types of lab tests ordered included urinalysis (n=77), malaria tests (n=59), and blood glucose tests (n=24). For the 209 total lab tests ordered, 35 diagnoses were made (**Supplement Figure 1**). The proportion referred to a clinician was similar among those taking 3HP via facilitated DOT vs. facilitated SAT (p=0.22). Those who took 3HP by facilitated SAT were more likely to have laboratory testing ordered (46.3% vs. 33.1%, p=0.04), whereas those who took 3HP by facilitated DOT were more likely to be prescribed medications to treat adverse events (78.9% vs. 67.4%, p=0.04).

Among the 270 PLHIV reporting adverse events, 40 (14.8%) had treatment held temporarily and 14 (5.1%) had treatment permanently discontinued by a Mulago ISS clinician (<1% of PLHIV taking 3HP overall). Participants taking 3HP via SAT (28.4% vs 15.4%) and participants enrolled after the first six months of the study (22.1% vs 15.7%) were more likely to have 3HP treatment held or discontinued due to an adverse event (**Supplement Table 3**).

## DISCUSSION

Growing interest in the use of short-course TPT regimens such as 3HP for TB prevention requires better data on adverse events to inform planning for programmatic scale-up. In this pragmatic trial of 1,655 PLHIV taking 3HP in Kampala, Uganda with minimal loss to follow-up prior to treatment completion, the frequency of adverse events was low, with only 16% of participants experiencing any adverse event. Adverse events were effectively managed by routine healthcare providers, with no deaths, only 3% requiring treatment interruption, and <1% requiring treatment discontinuation. These data add to the existing body of literature demonstrating 3HP safety and tolerability as some of the first data to come from a high-burden, programmatic context, and may provide reassurance to programs planning for or considering short-course regimens as alternatives for TPT in similar settings.

The frequency of adverse events reported in this pragmatic trial were comparable to what has been documented in other recent studies of 3HP implementation in high burden settings (12,13), but was substantially lower than what was reported from a multi-country randomized trial of 3HP (14), in which 77% reported one or more symptoms at any time during treatment (11). However, data from the only participating African country in that trial included only a small number of participants (n=83). Additionally, treatment dosing/supervision, study follow-up, and adverse event management in that trial were managed by trained research staff. Here, we provide data from people taking 3HP in the context of routine HIV/AIDS care, which may be a more accurate reflection of what can be expected in other programmatic settings. In this context, although the majority of participants who reported an adverse event were prescribed medications for those symptoms, the burden of laboratory testing was relatively modest.

As expected, we found that adverse events were more commonly reported with DOT than with SAT. DOT provides more opportunities for health workers to directly ask about and screen for potential side effects. It is also possible that PLHIV had a lower threshold for reporting potential side effects when face-to-face with a health worker than when prompted to so via IVR phone calls. Given the similar treatment completion rates between DOT and SAT, it is unlikely serious adverse events were more likely to be reported with one vs. the other.

A strength of this study was its pragmatic nature, including management of adverse events by routine healthcare providers. We therefore provide some of the first real-world evidence of 3HP adverse event occurrence from a clinical setting in a high-burden country. Limitations include the lack of a formal cost analysis of adverse event management and the possibility that people included in our study were more carefully screened for adverse events than would typically occur in the absence of a research study. As such, while we may overestimate the frequency of adverse events, management of those events including decisions to withhold or discontinue 3HP, may have been more careful.

In conclusion, this analysis of a pragmatic implementation trial in a large HIV clinic in Kampala, Uganda, reaffirms the safety of 3HP, with only one in six participants reporting an adverse event and fewer than 1% requiring permanent treatment discontinuation. These data also confirm that routine clinical staff can effectively manage 3HP-related adverse events. These data can be useful to National TB Programs and other implementing partners as they plan resources to support broader roll-out of short-course TPT, including an understanding of the potential additional time required and/or training needed for healthcare staff to manage 3HP-related adverse events and the potential additional costs for anticipated laboratory tests ordered and medications prescribed associated with 3HP-related adverse events. Based on our findings, these additional resources and/or costs associated with 3HP-related adverse events are likely to be low. Future studies of the total cost of 3HP-related adverse event management could provide a more complete picture of the adverse event profile likely to be experienced during programmatic scale-up. Nonetheless, these pragmatic data indicate that 3HP can be delivered safely and effectively in routine settings and can help implementers plan resources for adverse event management.

## Author contributions

JLK: Formal analysis, Data curation, Writing – Original draft, Writing – Review and editing, Visualization, Project administration; AM: Data curation, Validation, Data curation, Writing – Review and editing, Project administration; HEA: Formal analysis, Data curation, Writing – Original draft, Writing – Review and editing; LA: Data curation, Writing – Review and editing; FW: Data curation, Writing – Review and editing; JN: Data curation, Writing – Review and editing; CAB: Project administration, Writing – Review and editing; NK: Conceptualization, Methodology, Supervision, Writing – Review and editing; AK: Conceptualization, Methodology, Investigation, Supervision, Writing – Review and editing; DWD: Conceptualization, Methodology, Investigation, Supervision, Project Administration, Writing – Review and editing; AC: Conceptualization, Methodology, Investigation, Supervision, Project Administration, Funding acquisition, Writing – Review and editing; FS: Conceptualization, Methodology, Investigation, Supervision, Project Administration, Resource, Writing – Review and editing.

## Acknowledgements

The authors thank the administration, staff, and patients at the Mulago ISS clinic for their time and participation, and the research administration of the Infectious Diseases Research Collaboration (IDRC) and Walimu.

## Data availability

Data not publicly available.

