## Supplement Table 1 for "Adverse events reported during weekly isoniazid-rifapentine (3HP) tuberculosis preventive treatment among people living with HIV in Uganda"

**Supplement Table 1.** Logistic regression results assessing demographic and clinical characteristics associated with reporting at least one adverse event among all study participants initiating 3HP (n=1,651).

| Characteristic | Unadjusted Odds ratio (OR), 95% Confidence Interval (CI) | p-value | Adjusted OR,  (95% CI) | p-value |
| --- | --- | --- | --- | --- |
| *Age* | 1.00 (0.98-1.01) | 0.79 | 1.00 (0.99-1.02) | 0.70 |
| *Sex* |  |  |  |  |
| Male | *Reference* |  | *Reference* |  |
| Female | 1.61 (1.19-2.18) | 0.002 | 1.45 (1.05-2.01) | 0.02 |
| *BMI category* |  |  |  |  |
| Underweight (<18.5) | *Reference* |  | *Reference* |  |
| Normal: 18.5-24.9 | 0.96 (0.44-2.09) | 0.92 | 1.02 (0.46-2.24) | 0.96 |
| Overweight: 25-29.9 | 1.13 (0.51-2.47) | 0.77 | 1.14 (0.52-2.53) | 0.74 |
| Obesity: ≥30 | 1.52 (0.69-3.36) | 0.30 | 1.46 (0.65-3.26) | 0.36 |
| *Delivery strategy* |  |  |  |  |
| SAT | *Reference* |  | *Reference* |  |
| DOT | 1.57 (1.20-2.06) | 0.001 | 1.59 (1.21-2.09) | 0.001 |
| *MPI category* |  |  |  |  |
| Not poor/not vulnerable to poverty | *Reference* |  | *Reference* |  |
| Vulnerable to multidimensional poverty | 1.05 (0.77-1.42) | 0.76 | 1.01 (0.75-1.38) | 0.93 |
| Multidimensionally poor/severely multidimensionally poor | 1.43 (1.03-1.99) | 0.03 | 1.40 (1.00-1.96) | 0.05 |
| *Prior TB* |  |  |  |  |
| No prior TB | *Reference* |  | *Reference* |  |
| Prior TB | 0.72 (0.50-1.04) | 0.08 | 0.81 (0.56-1.18) | 0.27 |
| *Years on ART* |  |  |  |  |
| ≥1 year on ART | *Reference* |  | *Reference* |  |
| <1 year on ART | 1.09 (0.64-1.88) | 0.75 | 1.20 (0.69-2.10) | 0.53 |

ART=antiretroviral therapy; BMI=body mass index; DOT=directly observed therapy; MPI=multidimensional poverty index; SAT=self-administered therapy; TB=tuberculosis
