## Supplement Table 2 for "Adverse events reported during weekly isoniazid-rifapentine (3HP) tuberculosis preventive treatment among people living with HIV in Uganda"

**Supplement Table 2.** Reported adverse events reclassified into categories based on the Common Terminology Criteria for Adverse Events (version 5.0).

| CTCAE category | Adverse Event specified |
| --- | --- |
| Cardiac | Chest pain Palpitations |
| Dermatologic | Rash Itching Other skin problem |
| Ear disorder | Hearing loss Pain in ears |
| Eye disorder | Swelling of eyes/around eyes |
| Gastrointestinal | Diarrhea Loss of appetite Increased appetite Abdominal pain Gastritis Nausea Constipation Bloating Ulcers Oral sores Other oral problem |
| General/Systemic | Fever Weakness Generalized pain Oedema of limbs/face Flu/cold symptoms |
| Hepatobiliary | Yellow eyes or skin (jaundice) |
| Infection | Genital infection Urinary tract infection Shingles Pulmonary infection Pelvic inflammatory disease |
| Nervous system | Dizziness Numbness/Tingling/Peripheral neuropathy Headache Stroke |
| Musculoskeletal | Joint pain Back pain Neck pain Chest pain |
| Psychiatric | Drowsiness Insomnia Changes in sleep Restlessness Hallucinations |
| Renal | Painful micturition Renal cysts Frequent urination Polyuria Dysuria |
| Reproductive | Breast swelling/pain Vaginal pain Menstrual changes Pregnancy |
| Respiratory | Exacerbated asthma |
| Vascular | Pulmonary embolism |
