## Supplement Table 3 for "Adverse events reported during weekly isoniazid-rifapentine (3HP) tuberculosis preventive treatment among people living with HIV in Uganda"

**Supplement Table 3.** Logistic regression results assessing demographic and clinical characteristics associated with treatment hold or stop among those with adverse events (n=270).

| Characteristic | Unadjusted Odds ratio (OR),  95% Confidence Interval (CI) | p-value | Adjusted OR, (95% CI) | p-value |
| --- | --- | --- | --- | --- |
| *Age* | 1.02 (0.99-1.06) | 0.18 | 1.03 (0.99-1.06) | 0.14 |
| *Sex* |  |  |  |  |
| Male | Reference |  | Reference |  |
| Female | 1.00 (0.50-2.01) | 1.00 | 1.07 (0.51-2.24) | 0.86 |
| *Delivery strategy* |  |  |  |  |
| DOT | Reference |  | Reference |  |
| SAT | 2.18 (1.19-3.99) | 0.01 | 2.39 (1.26-4.52) | 0.007 |
| *Enrollment period* |  |  |  |  |
| January-July 2022 | Reference |  | Reference |  |
| July-December 2021 | 0.33 (0.14-0.79) | 0.01 | 0.36 (0.15-0.88) | 0.03 |
| January-June 2021 | 0.48 (0.20-1.13) | 0.09 | 0.41 (0.17-1.00) | 0.05 |
| July-December 2020 | 0.36 (0.16-0.80) | 0.01 | 0.33 (0.14-0.74) | 0.008 |
