## Supplement Figure 1 for "Adverse events reported during weekly isoniazid-rifapentine (3HP) tuberculosis preventive treatment among people living with HIV in Uganda"

**Supplement Figure 1.** Most reported laboratory test types and diagnoses made based on test results among study participants taking 3HP.

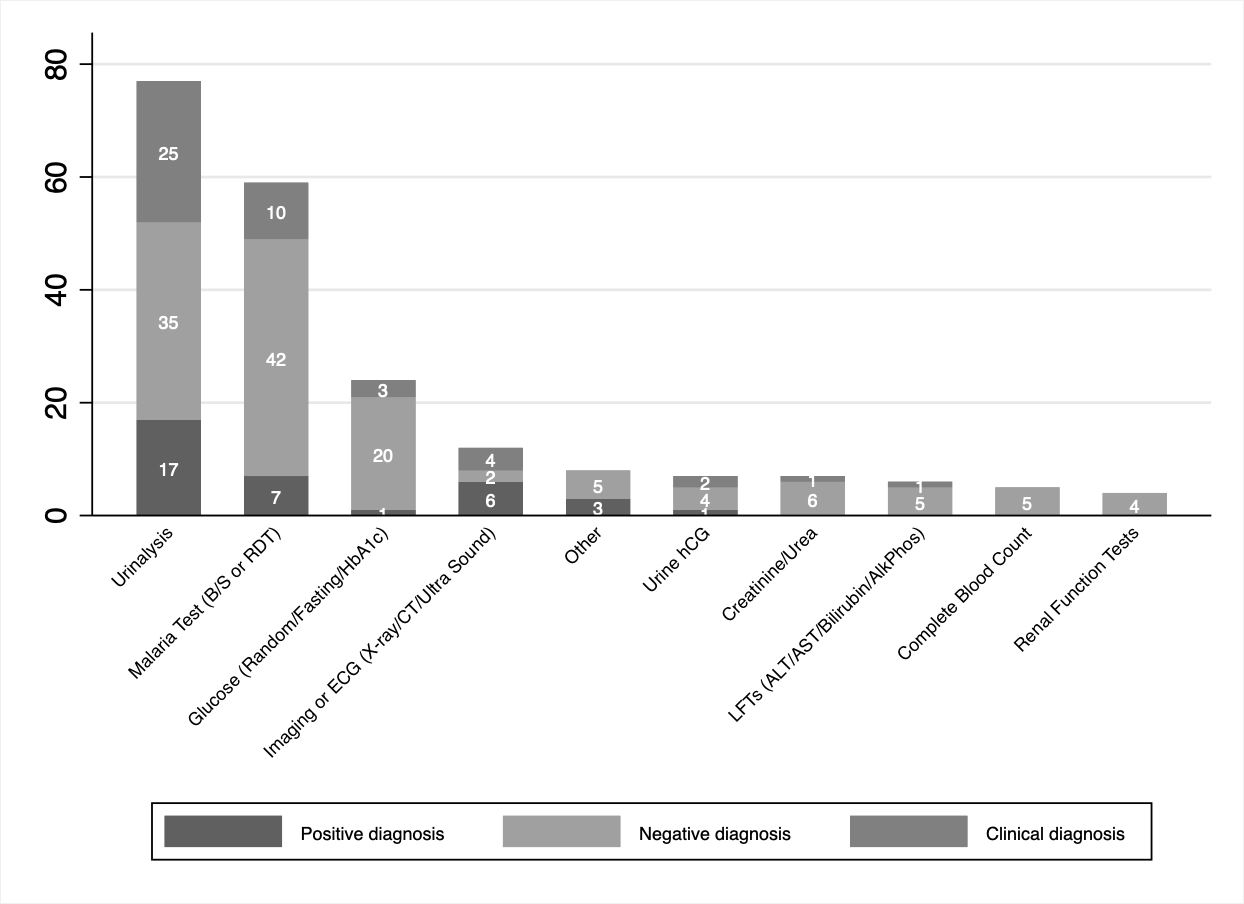

ALT=alanine transaminase, AST=aspartate transferase, B/S=blood smear, CT=computed tomography scan, ECG=echo cardiogram, hCG=human chorionic gonadotropin, LFT=liver function test, RDT=rapid diagnostic test
